# Genetic Susceptibility to Incisional Hernia Evaluation of Hernia Polygenic Risk Scores

**DOI:** 10.64898/2026.06.10.26355374

**Authors:** Andrew M. Pregnall, Margaret M. Hornick, Robyn B. Broach, Renae Judy, John DePaolo, Shuai Yuan, Michael G. Levin, John P. Fischer, Scott M. Damrauer, Heather Wachtel

**Affiliations:** Department of Surgery, University of Pennsylvania, Philadelphia, PA; Department of Surgery, MedStar Georgetown University Hospital, Washington, DC; Corporal Michael Crescenz VA Medical Center, Philadelphia, PA; Department of Medicine, University of Pennsylvania, Philadelphia, PA; Department of Genetics, University of Pennsylvania, Philadelphia, PA

**Keywords:** incisional hernia, polygenic risk scoring, risk modeling, Bayesian analysis

## Abstract

**Objectives:** Incisional hernia (IH) affects 13-30% of people after abdominal surgery, resulting in substantial morbidity and costs. While clinical risk factors have been studied extensively, genomic risk for IH is incompletely understood. We aimed to evaluate the impact of polygenic risk scores (PRS) on IH risk prediction.

**Methods:** We created and evaluated three PRS for abdominal hernia, ventral hernia and latent hernia susceptibility for prediction of IH in an institutional biobank. The primary outcome was defined as the diagnosis or repair of an IH based on ICD-9/10-CM/PCS and CPT codes. Clinical covariates included age, sex, body mass index (BMI), smoking status, index procedure type, and perioperative surgical site infection. A phenome-wide association study (PheWAS) was performed to assess clinical associations with increased PRS. We then tested the ability of the PRS to improve prediction for IH by modeling clinical covariates with and without PRS in patients who underwent abdominal surgery. Model performance was assessed using 10 iterations of 5-fold cross-validation to estimate Brier scores and area under the receiver operating characteristic curve (AUROC), which were compared using cross-model Bayesian analysis of variance.

**Results:** In 55,809 subjects, assessed PRS was significantly associated with incisional, umbilical, and ventral hernia on PheWAS, with 1.19 greater odds of developing IH per 1-SD increase in PRS (95% CI: 1.13-1.25, P < 0.001). Of 9,909 subjects who underwent qualifying abdominal surgery, 706 developed IH. In this cohort, the latent hernia susceptibility PRS was associated with a 16% increased hazard of developing IH per 1-SD increase (HR 1.16; 95% CI: 1.07-1.26; P < 0.001). Compared to a predictive model using clinical covariates (Brier score = 0.047, 95% CI: 0.046-0.048; AUROC = 0.660, 95% CI: 0.653-0.666), addition of the PRS showed similar Brier score and AUROC estimates (Brier score = 0.047, 95% CI: 0.046-0.048; AUROC: 0.667, 95% CI: 0.661-0.673) at five years. Cross-model Bayesian analysis demonstrated >99% probability of practical equivalence when trying to detect a difference of ≥ 0.02.

**Conclusion:** All three PRS for hernia were independently associated with IH, suggesting that genomic factors contribute significantly to IH development. However, none of the three PRS meaningfully improved clinical IH risk prediction in patients who underwent abdominal surgery. This suggests that clinical comorbidities and surgical techniques may be equally as important as genomic architecture.

## Introduction

Incisional hernia (IH) is a common complication following abdominal surgery affecting 10-20% of the general population and 30-50% of high risk patients and contributing to substantial surgical morbidity and population-level health burden. ^1–4^ IH significantly impacts patients’ quality of life including increased abdominal pain, poorer body image, rates of disability, and risk of incarceration.5 IH also poses in significant healthcare costs. In the U.S. alone there were an estimated 610,000 ventral hernia repairs resulting in $9.7 billion in expenditures in 2019. ^6,7^

Despite these efforts, IH remains a persistent and durable clinical problem, with a high recurrence rate after repair, making prevention the best treatment. Surgical techniques for hernia prevention have been extensively studied, including the role of operative approach, closure technique, suture material, and prophylactic mesh placement. Randomized clinical trials have suggested that a small bite or short stitch suture technique may result in lower odds of developing IH compared to large bite suture technique.^8–11^ Similarly, randomized clinical trials have suggested that primary placement of prophylactic mesh is more efficacious than non-mesh based approaches in high risk patient populations. ^1,2,12^ Although prophylactic mesh reconstruction has been demonstrated to be safe and effective, there remains concern about the broad use of mesh in patients undergoing abdominal surgery. ^13,14^ Therefore, identifying patients at highest risk of developing IH is important to guide clinical decision making.

Several clinical and operative risk factors for IH have been identified including smoking, increased body mass index (BMI), chronic obstructive pulmonary disease, abdominal aortic aneurysms, and abdominal operation type. ^1,2,15–17^ Although risk prediction models utilizing clinical variables have been developed, performance remains modest, and none have incorporated genetic effects. ^15,16,18,19^ Polygenic risk scores (PRS) sum the small genetic effects identified in genome-wide association studies into standardized scores and have been shown to predict complex traits. ^20–24^ In this study, we sought to evaluate whether addition of hernia PRS improved the calibration and discrimination of IH risk prediction.

## Methods

### Study population

This study was approved by the University of Pennsylvania Institutional Review Board (Protocol #24-1983). The primary analysis was performed in the Penn Medicine Biobank (PMBB), a genomic and precision medicine initiative that links genomic information to electronic health records of participants receiving care within the University of Pennsylvania Health System (UPHS). Currently, the PMBB contains 55,809 genotyped samples. To ascertain whether a PRS improved our ability to predict IH, we identified a cohort of PMBB participants who underwent abdominal surgery using International Classification of Disease (ICD) 9, ICD-10, and Clinical Procedural Terminology codes (see Supplementary Data). Index operations were defined as the first abdominal operation patients received within UPHS. Patients with a preexisting diagnosis of ventral hernia or whose index abdominal operation was a ventral or IH repair were excluded. The primary outcome was development of IH within five years of the index operation (see **Incisional Hernia Phenotyping**). Patients were censored at death, loss to follow-up, or repeat abdominal surgery within the five-year follow-up period.

### Development of hernia polygenic risk scores

We developed three hernia PRS using previously published summary statistics from a genome-wide association study meta-analysis of hernia subtypes across five global biobanks (see Table S1).25 First, a ventral hernia PRS (Ventral-PRS) to directly model the outcome of interest. Second, an abdominal hernia PRS (Abdominal-PRS) that leveraged a composite phenotype of ventral, inguinal, and umbilical hernia cases, motivated by the hypothesis that broader case ascertainment would increase genomic discovery and improve PRS performance. Finally, a latent hernia susceptibility factor PRS (PRS-Hernia) that modeled shared genetic architecture across hernia subtypes using genomic structural equation modeling (Genomic-SEM), representing a novel latent trait of general hernia susceptibility. Allele weights for each score were estimated using PRS-CSx, a Bayesian polygenic prediction method that employs a continuous shrinkage on SNP effect sizes and jointly models summary statistics from multiple ancestry groups to improve cross-ancestry prediction.26

### Calculation of hernia polygenic risk scores

Data collection procedures for PMBB genetic data have previously been described.27 As variant allele frequencies and linkage disequilibrium differs between genetic ancestry groups, we utilized pgsc_calc to calculate individual PRS. Briefly, pgsc_calc is a standardized, open-source pipeline that utilizes reference data from the 1000 Genomes Project and Human Genomes Diversity Project to calculate a genetic principal component space and distribution of polygenic risk scores.28 Within this space, PRS are modeled as a linear function of the first four genetic principal components, and their squared residuals are modeled as a function of the first four genetic principal components using gamma regression. Individual polygenic risk scores are then calculated using the equation:

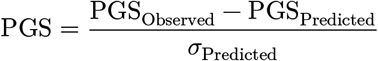

This methodology ensures that PRS values have a mean of 0 and standard deviation of 1 when considering the influence of population diversity.

### Statistical analysis

The discriminatory capacity of each PRS was first assessed by comparing score distributions between participants with and without prevalent hernia in the PMBB. A phenome-wide association study (PheWAS) was performed to characterize associations with elevated PRS across the phenome.

To evaluate whether PRS improved prediction of IH beyond established clinical risk factors, Cox proportional hazards models were constructed with age, sex, BMI, smoking status, surgical site infection (SSI), index procedure type, and PRS as covariates. Index procedures were classified as gynecological, vascular, hepatobiliary, colorectal, upper gastrointestinal, bariatric, urological, or transplant. 6.1% of participants were missing BMI values; missing values were imputed using the K-Nearest Neighbor algorithm (R package *VIM*), with all clinical covariates except PRS included as imputation predictors to avoid collinearity between imputed BMI values and the PRS. SSI was defined as a diagnosis of SSI in the 30 days preceding or following an index operation. Five-year IH probability was estimated from the Cox model using the R package *marginaleffects*.

Model discrimination was assessed using the time-dependent area under the receiver operating characteristic curve (AUROC) at one, three, and five years. Model calibration was assessed using the Brier score, calculated with the R package *yardstick*. To generate point estimates and 95% confidence intervals, five repeats of 10-fold cross-validation (50 total replicates) were performed. The incremental value of adding the PRS to the clinical model was assessed using a Bayesian approach. Specifically, the analysis of variance function in the R package *tidyposterior* was used to estimate the posterior distribution of the mean difference in AUROC and Brier scores between models, with 95% credible intervals reported. Practical equivalence was evaluated using a region of practical equivalence (ROPE) of 2%, with the probability of practical difference or equivalence reported for each model comparison.

### Reporting standards

This study followed Strengthening the Reporting of Observational Studies in Epidemiology (STROBE) reporting guidelines for cohort studies and the Polygenic Risk Score Standards Reporting Standards (PRS-RS) reporting guidelines. In the main text, we present the results from PRS-Hernia as the primary analysis. We include results from PRS-Ventral and PRS-Abdominal in the Supplementary Material. Results were consistent across PRS.

## Results

### Clinical and demographic characteristics of validation cohort

The overall study design is presented in Figure 1. Of 55,809 participants included, 11,623 had a diagnosis of any hernia and 2,185 had a diagnosis of ventral hernia. Among all participants, 11,114 individuals who underwent a qualifying abdominal surgery were identified. We excluded 1,119 individuals for having a pre-existing diagnosis of ventral/IH prior to their index abdominal operation and 86 individuals for having a ventral/IH repair as their index operation. The remaining 9,909 individuals were classified as the incident disease cohort. Of these, 706 developed IH within five years of their index abdominal operation; 5,109 did not develop ventral/IH within five years; 1,608 had a second abdominal operation within five years; 1,934 were lost to follow-up; and 552 were deceased within five years.

**Figure 1:**
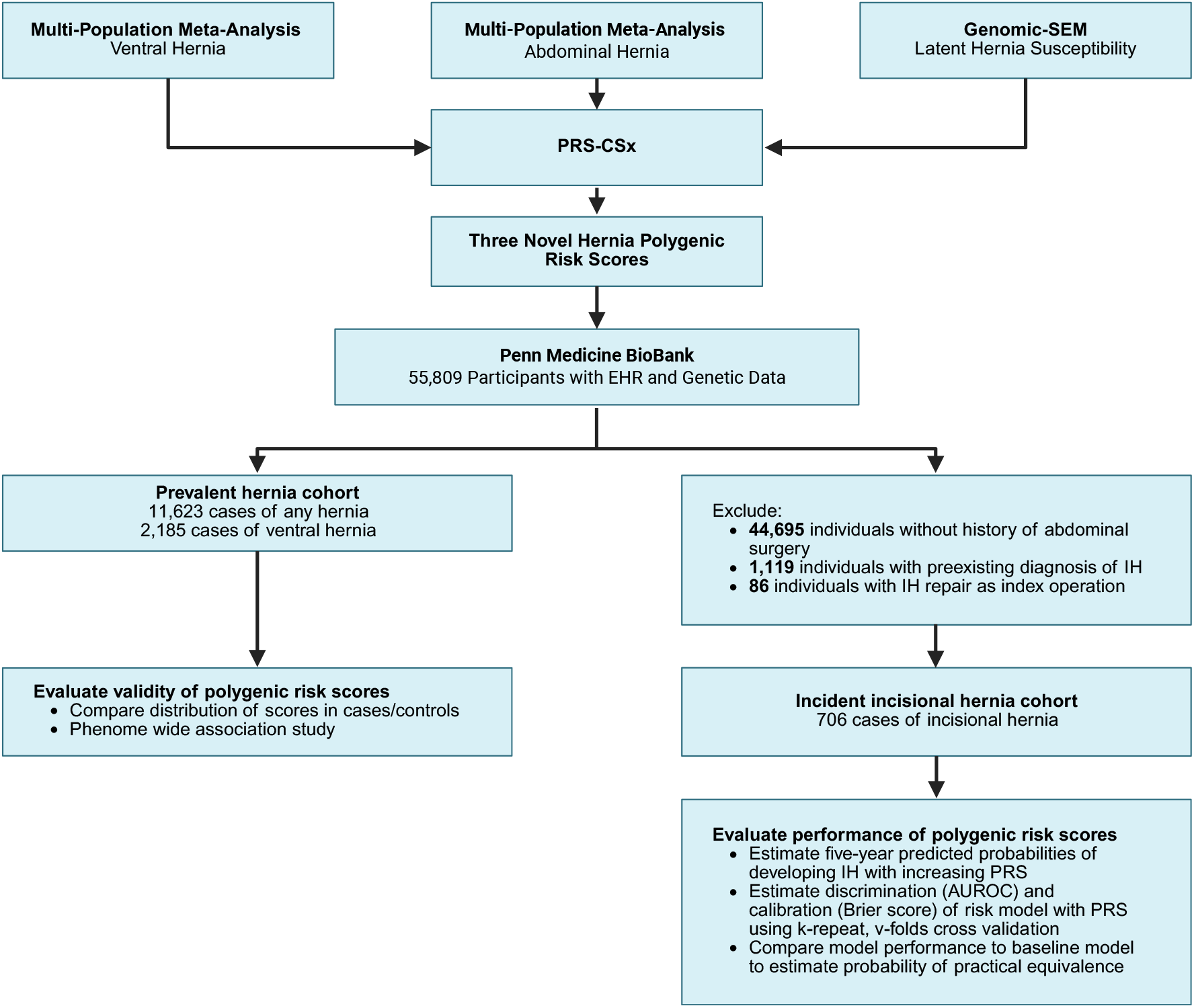
Overview of study design and cohort selection.

Compared to individuals who did not develop IH, individuals with IH were older (57.6 vs. 54.5 years; P < 0.001), more frequently genetically similar to the 1000 Genome European reference population (71.5% vs. 67.2%), more frequently underwent colorectal surgery as their index operation (32.6% vs. 17.8%), and had higher rates of SSI (8.4% vs. 2.8%; P < 0.001). BMI and smoking did not vary significantly by IH status. Demographic and clinical characteristics of the cohort are presented in Table 1.

**Table 1.**
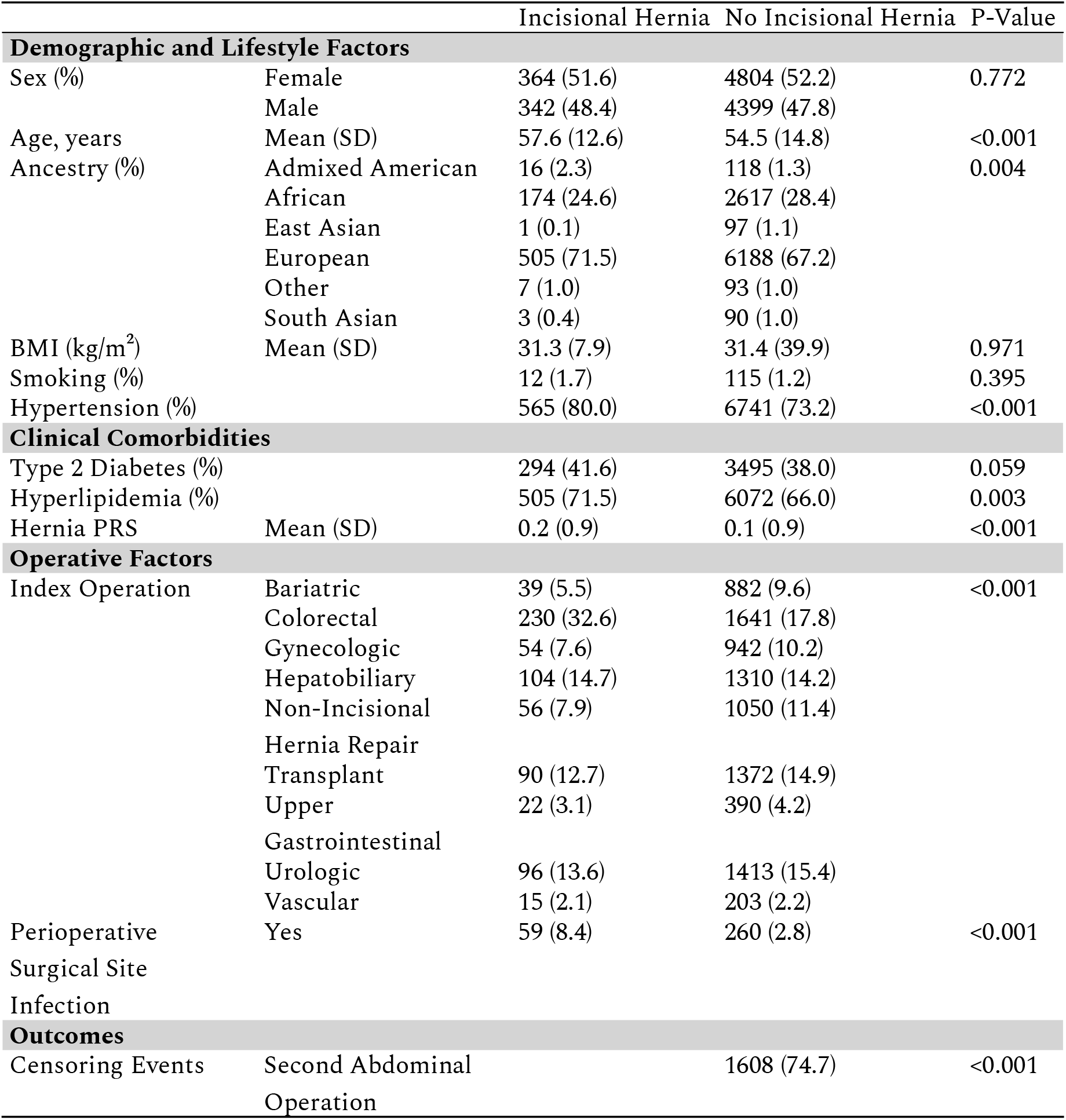
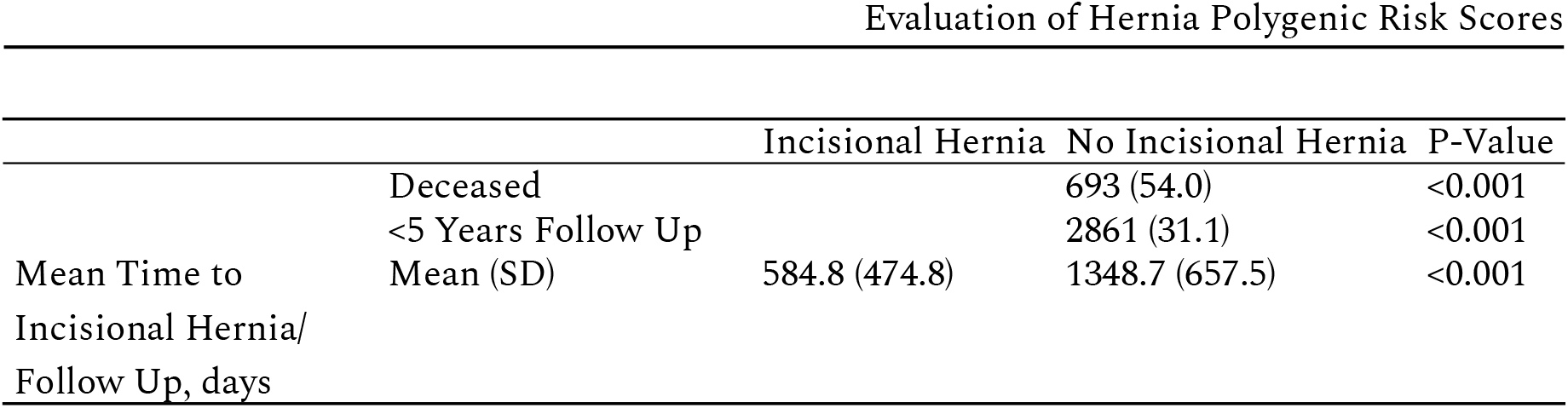
Clinical and Demographic Characteristics of Patients Undergoing Abdominal Surgery, Stratified by Incisional Hernia Status.

**Table 2.**
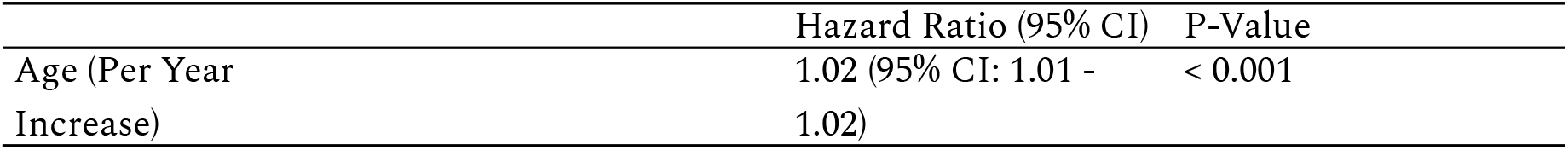
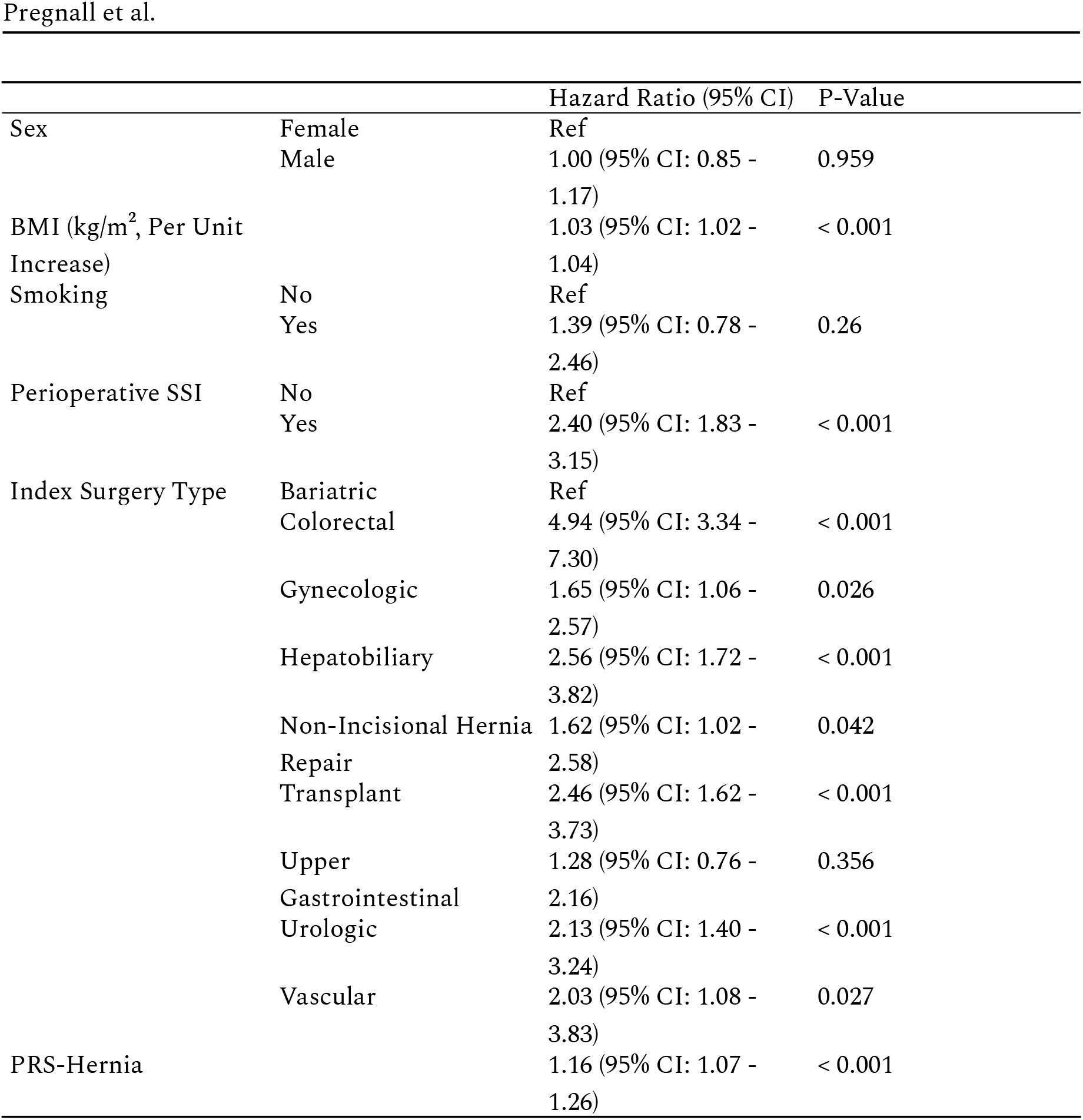
Association of PRS-Hernia with Incisional Hernia When Adjusting for Clinical Risk Factors.

|  | Hazard Ratio (95% CI) | P-Value |
| --- | --- | --- |
| Age (Per Year Increase) | 1.02 (95% CI: 1.01 - 1.02) | < 0.001 |

|  |  | Hazard Ratio (95% CI) | P-Value |
| --- | --- | --- | --- |
| Sex | Female | Ref |  |
|  | Male | 1.00 (95% CI: 0.85 - 1.17) | 0.959 |
| BMI (kg/m<sup>2</sup>, Per Unit Increase) |  | 1.03 (95% CI: 1.02 - 1.04) | < 0.001 |
| Smoking | No | Ref |  |
|  | Yes | 1.39 (95% CI: 0.78 - 2.46) | 0.26 |
| Perioperative SSI | No | Ref |  |
|  | Yes | 2.40 (95% CI: 1.83 - 3.15) | < 0.001 |
| Index Surgery Type | Bariatric | Ref |  |
|  | Colorectal | 4.94 (95% CI: 3.34 - 7.30) | < 0.001 |
|  | Gynecologic | 1.65 (95% CI: 1.06 - 2.57) | 0.026 |
|  | Hepatobiliary | 2.56 (95% CI: 1.72 - 3.82) | < 0.001 |
|  | Non-Incisional Hernia Repair | 1.62 (95% CI: 1.02 - 2.58) | 0.042 |
|  | Transplant | 2.46 (95% CI: 1.62 - 3.73) | < 0.001 |
|  | Upper Gastrointestinal | 1.28 (95% CI: 0.76 - 2.16) | 0.356 |
|  | Urologic | 2.13 (95% CI: 1.40 - 3.24) | < 0.001 |
|  | Vascular | 2.03 (95% CI: 1.08 - 3.83) | 0.027 |
| PRS-Hernia |  | 1.16 (95% CI: 1.07 - 1.26) | < 0.001 |

### Association of hernia polygenic risk score with prevalent hernia in a diverse institutional biobank

We sought to assess the polygenic risk scores in an independent cohort of subjects in the PMBB. Individuals with a diagnosis of any hernia had significantly higher mean PRS-Hernia compared to individuals without any hernia diagnosis (0.1 vs. 0.01; P < 0.001), and individuals with a diagnosis of ventral hernia had a significantly higher mean PRS-Hernia compared to individuals without a ventral hernia diagnosis (0.15 vs. 0.02: P < 0.001). This finding was consistent across polygenic risk scores (see Table S2).

To characterize the strength of the PRS-Hernia, we compared its association with prevalent hernia in the PMBB. PRS-Hernia was associated with 1.16 greater odds of developing ventral hernia per 1-SD increase (95% CI: 1.11-1.21, *P* < 0.001). The strengths of association were consistent when stratifying by genetically similar population group; however, less precise estimates were observed in non-European population groups (see Table S3). Finally, we performed an unbiased PheWAS between the PRS-Hernia and prevalent disease in PMBB. PRS-Hernia demonstrated significant associations with phenotypes including morbid obesity (OR: 1.11; 95% CI: 1.07-1.14; P < 0.001), abdominal pain (OR: 1.05; 95% CI: 1.03-1.07; P < 0.001), obstructive sleep apnea (OR: 1.05; 95% CI: 1.03-1.07; P < 0.001), gastroesophageal reflux disease (OR: 1.04; 95% CI: 1.02-1.06; P < 0.001), and diverticulosis (OR: 1.05; 95% CI: 1.03-1.08; P < 0.001), with OR expressed as per 1-SD increase in PRS-Hernia.

### Adjusted association of hernia polygenic risk scores with incisional hernia

To determine if the latent-factor hernia polygenic risk score was independently associated with incident IH, we performed a multivariable survival analysis using the Cox proportional hazards model, adjusting for age, sex, BMI, smoking, index surgery type, SSI, and PRS-Hernia. PRS-Hernia was associated with 16% increased hazard of developing IH per 1-SD increase (Hazard Ratio: 1.16; 95% CI: 1.07-1.26; P < 0.001). There was no evidence of interaction between PRS-Hernia and index surgery type (P = 0.791, Analysis of Deviance) or PRS-Hernia and BMI (P = 0.066, Analysis of Deviance).

We estimated counterfactual predicted probabilities of developing IH at five years following an index abdominal operation for each risk factor (see Figure 2). Compared to individuals with a PRS-Hernia of 0 (Predicted Probability = 9.1%; 95% CI: 6.1%-12.1%), individuals with a PRS-Hernia two standard deviations above the scaled population average had a five-year predicted probability developing IH of 12% (95% CI: 7.8%-16.1%), while individuals two standard deviations below the average had a predicted probability of 6.8% (95% CI: 4.3%-9.4%). Across BMI categories, predicted IH probability at five years ranged from 6.9% (95% CI: 4.8%-9.0%) among underweight individuals to 9.9% (95% CI: 6.5%-13.2%) among obese individuals. Index surgery type was the strongest predictor of five-year IH risk: colorectal surgery was associated with the highest predicted probability of developing IH (17.6%; 95% CI: 11.4%–23.9%), whereas bariatric surgery was associated with the lowest (3.9%; 95% CI: 3.6%–4.2%). Finally, individuals with an SSI had a markedly higher five-year predicted probability of developing IH (19.5%; 95% CI: 12.2%-26.7%) compared to those without SSI (8.8%; 95% CI: 5.9%-11.7%).

**Figure 2:**
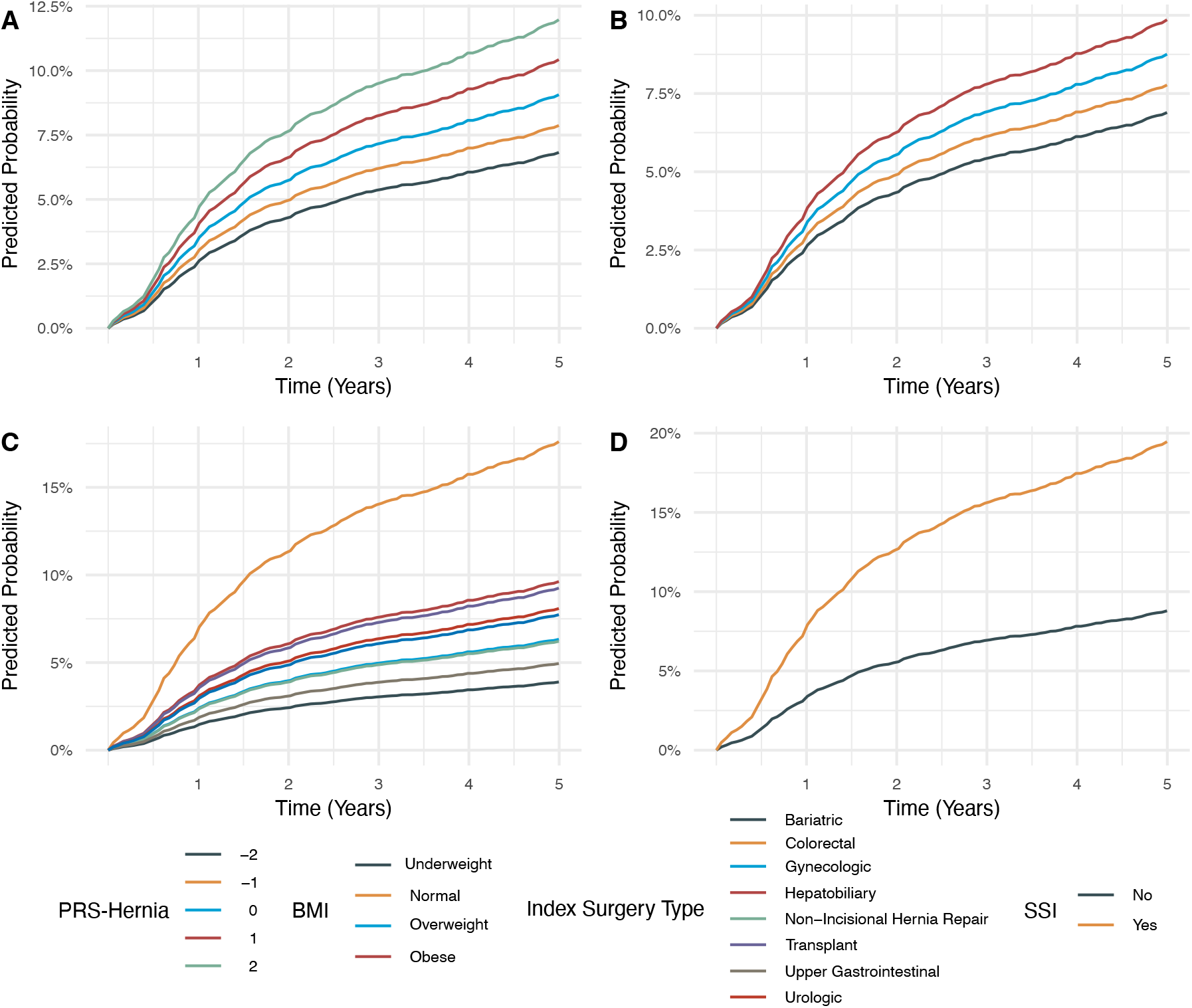
Predicted probabilities of developing incisional hernia by key covariates. Kaplan-Meier-style curves show model-estimated probability of remaining hernia-free over five years following index abdominal surgery, stratified by (A) PRS-Hernia, (B) BMI category, (C) index surgery type, and (D) surgical site infection status.

**Figure 3:**
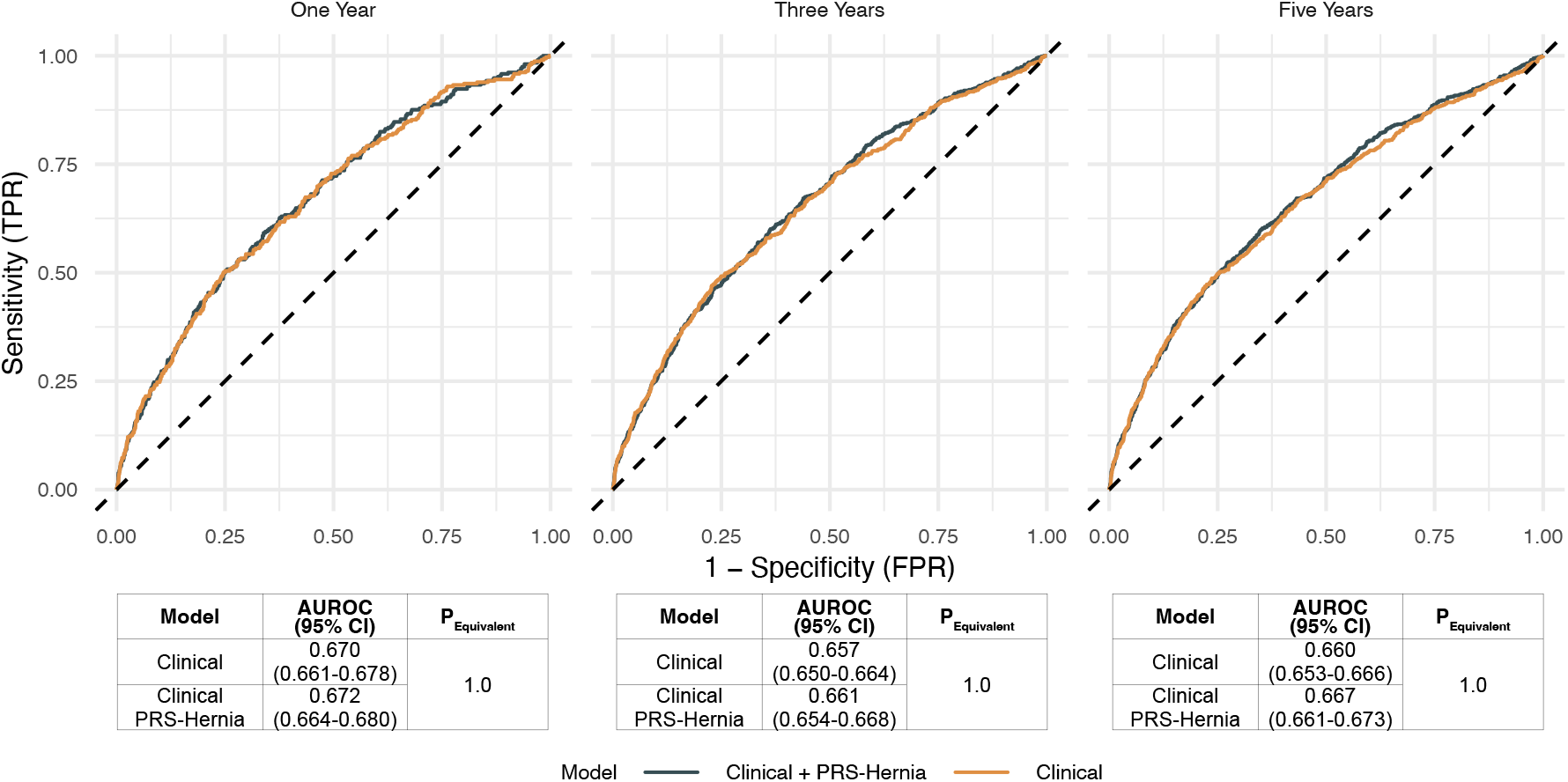
Time-dependent receiver operating characteristic curves for incisional hernia prediction at one, three, and five years. ROC curves compare discriminative performance of a clinical-only model versus a model augmented with PRS-Hernia (Clinical + PRS-Hernia) at each time horizon. Tables below each panel report the Area Under the ROC Curve (AUROC) with 95% confidence intervals and the probability of equivalence (P-Equivalent) between models. The two models demonstrated near-identical discrimination across all time points (AUROC ∼0.67–0.68), with no meaningful improvement conferred by the addition of PRS-Hernia.

### Assessing the Utility of a Hernia Polygenic Risk Score in Post-Operative Hernia Prediction Models

To determine if PRS-Hernia could improve IH risk stratification for individuals undergoing abdominal surgery, we integrated PRS-Hernia into a clinical risk prediction model and tested it in the incident disease cohort. To assess model calibration, cross validation was performed to generate Brier scores for a baseline Age + Sex + BMI + Smoking Status + Index Surgery + SSI model and an integrated Age + Sex + BMI + Smoking Status + Index Surgery + SSI + PRS-Hernia model. Compared to the baseline model (Brier score = 0.047; 95% CI: 0.046-0.048), the integrated model had similar Brier score estimates (Brier score = 0.047; 95% CI: 0.046-0.048). To formally determine if the addition of the PRS-Hernia improved model calibration, we employed cross-model Bayesian ANOVA analysis. The integrated and clinical models demonstrated a high probability of being practically equivalent in their Brier scores (Mean Difference: < 0.001; 95% CI: < 0.001 - < 0.001; Probability of Equivalence: 100%; see Table S6).

To assess model discrimination, cross validation was performed to generate AUROC estimates for the risk prediction models at one, three, and five-year time horizons. Compared to the baseline model at five years (AUROC = 0.660; 95% CI: 0.650-0.664), the integrated model had similar AUROC estimates (AUROC: 0.661; 95% CI: 0.654-0.668), and cross-model Bayesian ANOVA analysis demonstrated a high likelihood of model equivalence (Mean Difference: 0.007; 95% CI: 0.001-0.012; Probability of Equivalence: >99%). These findings were robust across polygenic risk scores (see Figure S1 and Figure S2).

## Discussion

In this study, we developed multiple hernia polygenic risk scores using diverse, multi-population data from five global biobanks. To our knowledge, this is the first study that reports the develop-ment of hernia polygenic risk score using multi-population data. ^22,29,30^ Furthermore, it is the first study to examine the utility of a hernia polygenic risk score in predicting IH. ^15,16,18,19^

The polygenic risk scores were significantly associated with prevalent hernia in an independent, diverse institutional biobank. Notably, PRS-Hernia, which captures shared genetic liability across hernia subtypes, demonstrated associations across multiple hernia types, suggesting broad utility across the hernia phenotypic spectrum. In addition, the magnitude of association between PRS-Hernia and IH exceeded that of BMI which is the most widely included clinical risk factor for hernia, underscoring the potential relevance of inherited genetic predisposition in hernia susceptibility which is estimated to be 10-20% for non-diaphragmatic hernia subtypes.25 These findings demonstrate the value of integrating genetic risk across populations and support the potential clinical impact of polygenic risk score stratification for predicting IH.

While the hernia PRS were significantly associated with IH development, the PRS did not significantly improve IH risk stratification compared to clinical factors alone in a large cohort of patients undergoing abdominal surgeries. These findings highlight the difference between identifying statistical associations and improving predictive performance. ^31,32^ While numerous associations have been identified between IH and surgical factors, clinical phenotypes, and imaging-derived biomarkers, ^33^ challenges remain in translating these findings into rigorous risk prediction models with strong discriminative capacity. ^15,16^ Our finding is consistent with studies in cardiovascular literature that have demonstrated polygenic risk scores do not improve predictive performance of clinical risk prediction models. ^34,35^ This may represent modest incremental discrimination in baseline models which are already well calibrated, or alternatively may be due to overlap between clinical conditions and genomic contributions.

The vision of precision medicine initiatives is to tailor diagnosis and treatment options to individual patients. In the case of IH, this means identifying patients that would benefit most from preventative measures such as placement of prophylactic mesh reinforcement at the time of surgery. While complication rates from prophylactic mesh placement are low, concern still remains about their broad usage. ^13,14^ Therefore, developing strategies for patient stratification is an important goal. Previous work has shown that a risk prediction model would need a discriminatory capacity of 80% to influence surgical decision making. ^36^ While hernia polygenic risk scores are not sufficient to independently realize this vision, this limitation is similarly shared by current clinical risk predictors. For example, while strongly associated with hernia formation, BMI alone does not meaningfully improve IH risk models. This reflects a broader reality that few clinical or genetic variables are sufficient to independently improve risk stratification models for multifactorial phenotypes like IH. However, our data also shows that incorporation of multiple variables improved the AUROC from 50 to 70%, demonstrating that PRS and other variables offer incremental improvements to risk prediction models when utilized together. Therefore, incorporation of PRS along with other data such as imaging-derived phenotypes may allow for improved patient stratification by capturing factors such as baseline genetic risk. Further investigation is needed in this area.

Our study also found that non-modifiable risk factors such as index surgery type and SSI are strongly associated with IH development. Consistent with prior work, colorectal surgery and SSI had the highest association with IH development. ^15,16^ The dominance of perioperative factors over the PRS in our predicted probability models suggests that IH formation following abdominal surgery is driven primarily by procedural context rather than baseline genetic predisposition. However, there are other contexts in which PRS may be useful. Prior work has demonstrated associations between an inguinal hernia polygenic risk score and hernia severity and recurrence.37 Future studies could assess the utility of a polygenic risk score in predicting recurrent IH following abdominal surgery or IH severity. In addition, previous studies have shown that polygenic risk scores can be used to motivate behavioral change.38 Further studies could examine the role of hernia polygenic risk scores in motivating patients to change modifiable risk factors such as BMI and smoking.

## Limitations

This study has several limitations. First, some patients in the risk prediction cohort may have previous diagnoses or repairs of ventral/IH at outside hospitals not captured in the PMBB. Our study assumes these patients—who otherwise would have been excluded from analysis—do not have a history of ventral/IH diagnosis or repair. Similarly, some patients in the risk prediction cohort may have had qualifying abdominal surgeries at outside hospital systems prior to their enrollment in the PMBB. As an increasing number of abdominal surgeries is a known risk factor for IH, we define patient’s index procedure as their first abdominal operation at our institution and therefore may under-capture patients who had prior surgery at other institutions.

## Conclusion

We found that polygenic risk scores developed in abdominal hernia, ventral hernia and latent hernia susceptibility were significantly associated with IH development in an independent validation cohort, suggesting that there is a strong genetic component to the development of IH. Although PRS was more predictive than BMI alone, the addition of a hernia PRS to a clinical IH risk prediction model did not significantly improve the model’s discriminative capacity. This nuanced finding suggests that genomic risk scores have a role not as standalone decision tools, but as components of multi-modal risk models.

## Data Availability

Allele weights for the polygenic risk scores will be deposited in the PGS Catalog upon publication. Access to Penn Medicine Biobank is available upon application.  

## Penn Medicine BioBank

We acknowledge the Penn Medicine BioBank (PMBB) for providing data and thank the patient-participants of Penn Medicine who consented to participate in this research program. We would also like to thank the PMBB team and Regeneron Genetics Center for providing genetic variant data for analysis. The PMBB is approved under IRB protocol number 813913 and supported by Perelman School of Medicine at University of Pennsylvania, a gift from the Smilow family, and the National Center for Advancing Translational Sciences of the National Institutes of Health under CTSA award number UL1TR001878.

## Supplementary Material

### Incisional Hernia Phenotyping

Incisional hernia was ascertained using administrative and billing diagnosis and procedure codes from the electronic health record, including ICD-9-CM, ICD-10-CM, ICD-9-PCS, ICD-10-PCS, and Current Procedural Terminology (CPT) codes. Code lists used to define ventral/incisional hernia diagnosis and repair are provided in the Supplementary Data.

Patients were excluded from the at-risk cohort if they had evidence of a preexisting ventral or incisional hernia, defined as a ventral/incisional hernia diagnosis code occurring on or before their index abdominal operation. Patients were also excluded if the index operation was a ventral or incisional hernia repair, defined by the presence of any ventral/incisional hernia repair code on the date of their index operation. Primary ventral hernia codes could not be reliably distinguished from incisional hernia codes due to overlap in coding practices, subsequently, ventral and incisional hernia codes were analyzed together. Patients with umbilical hernia diagnosis or repair alone were not excluded.

The primary outcome was postoperative ventral/incisional hernia, defined as the first occurrence after the index operation of either a ventral/incisional hernia diagnosis code or a ventral/ incisional hernia repair code. Because ICD-10-PCS and newer CPT repair codes classify hernia repair by size rather than type, repair codes that could represent ventral/incisional or umbilical hernia repair were required to have a concurrent ventral/incisional hernia diagnosis code to be classified as the primary outcome.

Postoperative ventral/incisional hernia was defined as the first occurrence of the following:

- An ICD-9-CM diagnostic code for ventral/incisional hernia
- An ICD-10-CM diagnostic code ventral/incisional hernia
- An ICD-9-PCS repair code for ventral/incisional hernia
- A CPT repair code specific for ventral/incisional hernia
- An ICD-10-PCS repair code for ventral/incisional/umbilical hernia repair with an ICD-10-CM diagnostic code for ventral/incisional hernia
- A CPT repair code for ventral/incisional/umbilical hernia repair with either an ICD-9-CM or ICD-10-CM diagnostic code for ventral/incisional hernia

Outcome status was assessed within a five-year follow-up window from the index operation date. Patients were censored at the earliest of death, a subsequent non-hernia abdominal operation, date of last follow-up if less than five years after the index operation, or five years after the index operation.

---

**Table S1:**
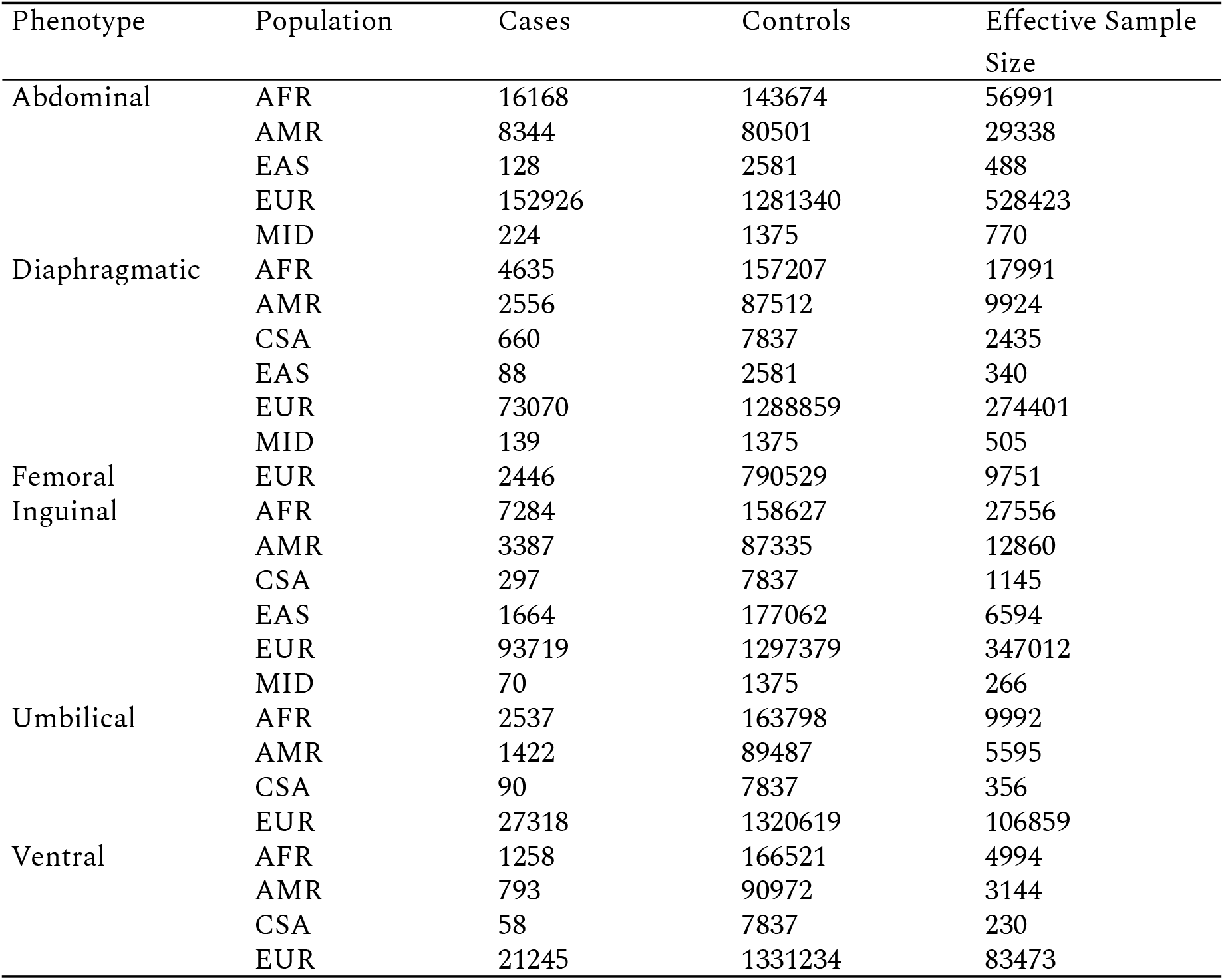
Summary of Number of Cases, Controls, and Effective Sample Size for Input Summary Statistics, Stratified by Phenotype and Population.

**Table S2:** Distribution of hernia polygenic risk scores, stratified by hernia subtype.

| Phenotype | Polygenic Risk<br>Score | Mean PRS Cases | Mean PRS<br>Controls | P-Value |
| --- | --- | --- | --- | --- |
| Any Hernia | Abdominal PRS | 0.13 | 0.02 | < 0.001 |
|  | Ventral PRS | 0.06 | 0.02 | < 0.001 |
| Ventral Hernia | Abdominal PRS | 0.18 | 0.04 | < 0.001 |
|  | Ventral PRS | 0.11 | 0.03 | < 0.001 |

**Table S3:** Clinical and Demographic Characteristics of Patients Undergoing Abdominal Surgery, Stratified by Incisional Hernia Status.

| Polygenic Risk Score | Ancestry | Odds Ratio (95% CI) | P-Value |
| --- | --- | --- | --- |
| PRSCSx_CommonFactor | EUR | 1.18 (1.11 to 1.25) | < 0.001 |
|  | AFR | 1.07 (0.99 to 1.17) | 0.087 |
|  | SAS | 1.34 (0.84 to 2.15) | 0.219 |
|  | EAS | 1.07 (0.36 to 3.35) | 0.91 |
|  | AMR | 1.45 (1.03 to 2.06) | 0.035 |
|  | Other | 1.07 (0.59 to 1.93) | 0.823 |
| PRSCSx_Abdominal | EUR | 1.17 (1.11 to 1.23) | < 0.001 |
|  | AFR | 1.09 (1.01 to 1.18) | 0.029 |
|  | SAS | 1.79 (1.09 to 2.97) | 0.023 |
|  | EAS | 0.41 (0.13 to 1.23) | 0.116 |
|  | AMR | 1.36 (0.96 to 1.92) | 0.081 |
|  | Other | 0.93 (0.51 to 1.7) | 0.82 |
| PRSCSx_Ventral | EUR | 1.1 (1.04 to 1.16) | < 0.001 |
|  | AFR | 1.02 (0.94 to 1.1) | 0.707 |
|  | SAS | 1.66 (1.00 to 2.77) | 0.051 |
|  | EAS | 0.83 (0.29 to 2.36) | 0.731 |
|  | AMR | 1.26 (0.90 to 1.77) | 0.181 |
|  | Other | 1.49 (0.83 to 2.73) | 0.188 |

**Table S4:** Association of PRS-Ventral with Incisional Hernia When Adjusting for Clinical Risk Factors.

|  |  | Hazard Ratio (95% CI) | P-Value |
| --- | --- | --- | --- |
| Age (Per Year Increase) |  | 1.02 (95% CI: 1.01 - 1.02) | < 0.001 |
| Sex | Female | Ref |  |
|  | Male | 0.99 (95% CI: 0.84 - 1.16) | 0.897 |
| BMI (kg/m<sup>2</sup>, Per Unit Increase) |  | 1.03 (95% CI: 1.02 - 1.04) | < 0.001 |
| Smoking | No | Ref |  |
|  | Yes | 1.38 (95% CI: 0.78 - 2.45) | 0.266 |
| Perioperative SSI | No | Ref |  |
|  | Yes | 2.42 (95% CI: 1.84 - 3.17) | < 0.001 |
| Index Surgery Type | Bariatric | Ref |  |
|  | Colorectal | 4.93 (95% CI: 3.33 - 7.28) | < 0.001 |
|  | Gynecologic | 1.63 (95% CI: 1.05 - 2.54) | 0.029 |
|  | Hepatobiliary | 2.57 (95% CI: 1.72 - 3.83) | < 0.001 |
|  | Non-Incisional Hernia Repair | 1.64 (95% CI: 1.03 - 2.61) | 0.037 |
|  | Transplant | 2.46 (95% CI: 1.62 - 3.74) | < 0.001 |
|  | Upper Gastrointestinal | 1.30 (95% CI: 0.77 - 2.19) | 0.328 |
|  | Urologic | 2.13 (95% CI: 1.40 - 3.24) | < 0.001 |
|  | Vascular | 2.04 (95% CI: 1.08 - 3.83) | 0.028 |
| PRS-Ventral |  | 1.10 (95% CI: 1.01 - 1.18) | 0.019 |

**Table S5:** Association of PRS-Abdominal with Incisional Hernia When Adjusting for Clinical Risk Factors.

|  |  | Hazard Ratio (95% CI) | P-Value |
| --- | --- | --- | --- |
| Age (Per Year Increase) |  | 1.02 (95% CI: 1.01 - 1.02) | < 0.001 |
| Sex | Female | Ref |  |
|  | Male | 1.00 (95% CI: 0.85 - 1.18) | 0.995 |
| BMI (kg/m<sup>2</sup>, Per Unit Increase) |  | 1.03 (95% CI: 1.02 - 1.04) | < 0.001 |
| Smoking | No | Ref |  |
|  | Yes | 1.37 (95% CI: 0.78 - 2.43) | 0.276 |
| Perioperative SSI | No |  |  |
|  | Yes | 2.42 (95% CI: 1.85 - 3.18) | < 0.001 |
| Index Surgery Type | Bariatric |  |  |
|  | Colorectal | 4.94 (95% CI: 3.34 - 7.29) | < 0.001 |
|  | Gynecologic | 1.65 (95% CI: 1.06 - 2.57) | 0.026 |
|  | Hepatobiliary | 2.58 (95% CI: 1.73 - 3.85) | < 0.001 |
|  | Non-Incisional Hernia Repair | 1.62 (95% CI: 1.01 - 2.57) | 0.043 |
|  | Transplant | 2.46 (95% CI: 1.62 - 3.74) | < 0.001 |
|  | Upper Gastrointestinal | 1.29 (95% CI: 0.77 - 2.19) | 0.334 |
|  | Urologic | 2.13 (95% CI: 1.41 - 3.24) | < 0.001 |
|  | Vascular | 2.03 (95% CI: 1.08 - 3.83) | 0.028 |
| PRS-Abdominal |  | 1.12 (95% CI: 1.04 - 1.20) | 0.003 |

**Table S6:** Brier score estimates and probability of equivalence.

| **Model** | **Brier Score** | **Brier Score 95% CI** | **Model Difference** | **Model Difference 95% CI** | **Probability of Equivalence** |
| --- | --- | --- | --- | --- | --- |
| Clinical | 0.047 | 0.046-0.048 | < 0.001 | <0.001 - < 0.001 | 100% |
| Clinical + PRS-Hernia | 0.047 | 0.046-0.048 |  |  |  |
| Clinical | 0.047 | 0.046-0.048 | < 0.001 | <0.001 - < 0.001 | 100% |
| Clinical + PRS-Ventral | 0.047 | 0.046-0.048 |  |  |  |
| Clinical | 0.047 | 0.046-0.048 | < 0.001 | <0.001 - < 0.001 | 100% |
| Clinical + PRS-Abdominal | 0.047 | 0.046-0.048 |  |  |  |

**Figure S1:**
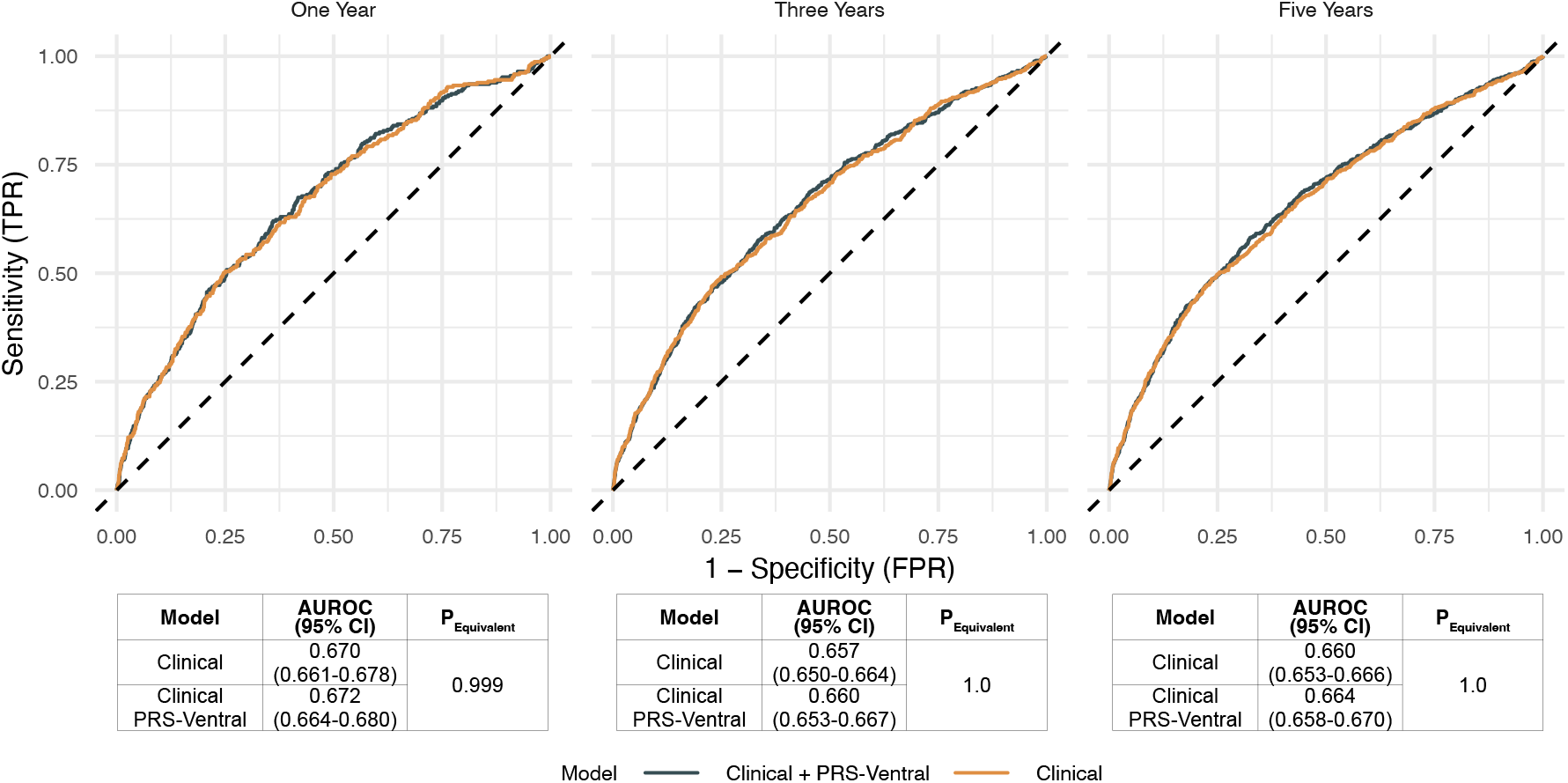
Time-dependent receiver operating characteristic curves for incisional hernia prediction at one, three, and five years. ROC curves compare discriminative performance of a clinical-only model versus a model augmented with PRS-Ventral (Clinical + PRS-Ventral) at each time horizon. Tables below each panel report the Area Under the ROC Curve (AUROC) with 95% confidence intervals and the probability of equivalence (P-Equivalent) between models. The two models demonstrated near-identical discrimination across all time points (AUROC ∼0.67–0.68), with no meaningful improvement conferred by the addition of PRS-Ventral.

**Figure S2:**
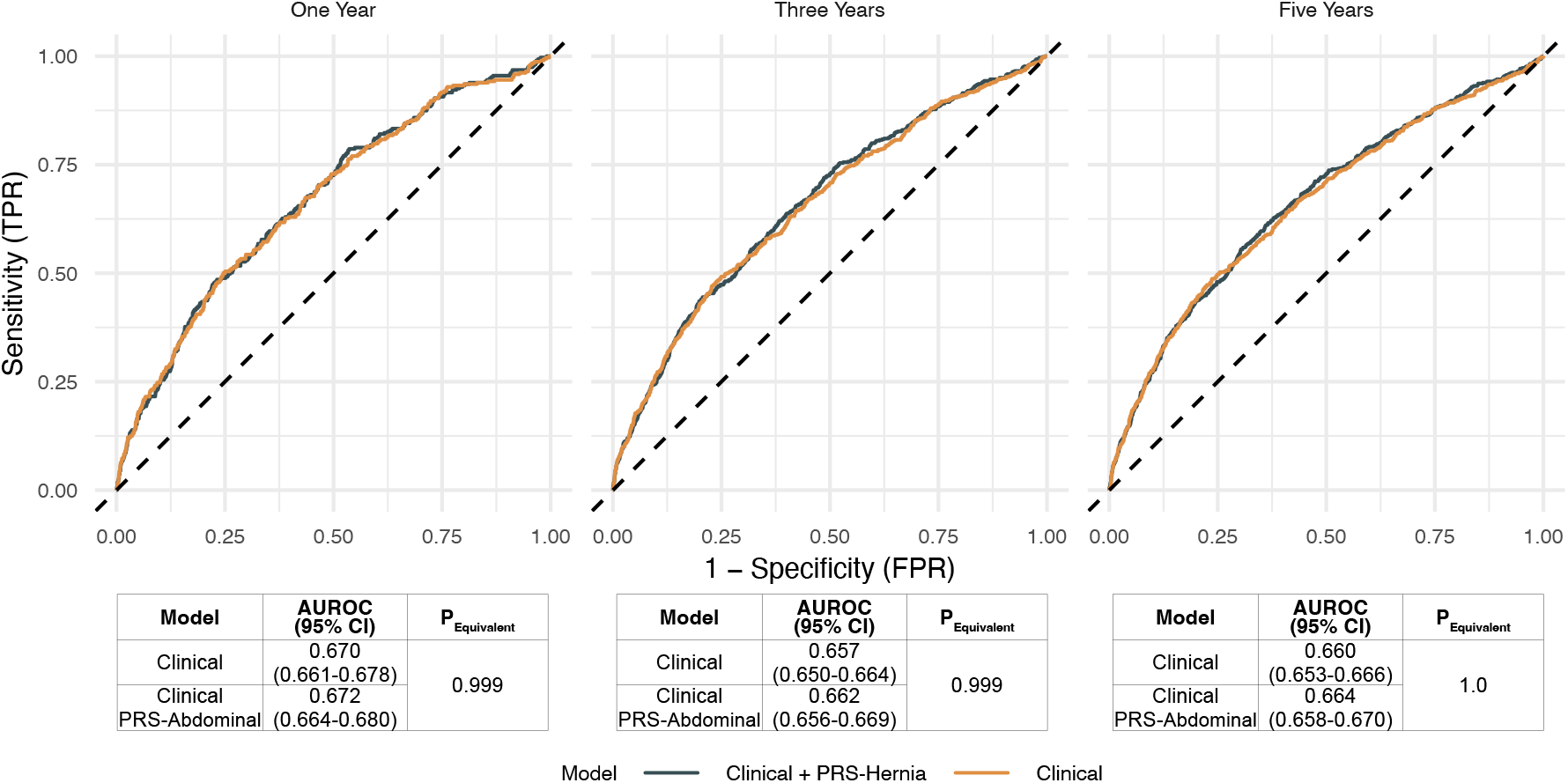
Time-dependent receiver operating characteristic curves for incisional hernia prediction at one, three, and five years. ROC curves compare discriminative performance of a clinical-only model versus a model augmented with PRS-Abdominal (Clinical + PRS-Abdominal) at each time horizon. Tables below each panel report the Area Under the ROC Curve (AUROC) with 95% confidence intervals and the probability of equivalence (P-Equivalent) between models. The two models demonstrated near-identical discrimination across all time points (AUROC ∼0.67–0.68), with no meaningful improvement conferred by the addition of PRS-Abdominal.

## References

1. Van Den Dop LM, Sneiders D, Yurtkap Y, et al. Prevention of incisional hernia with prophylactic onlay and sublay mesh reinforcement vs. primary suture only in midline laparotomies (PRIMA): long-term outcomes of a multicentre, double-blind, randomised controlled trial. The Lancet Regional Health - Europe. 2024;36:100787. doi:10.1016/j.lanepe.2023.100787

2. Jairam AP, Timmermans L, Eker HH, et al. Prevention of incisional hernia with prophylactic onlay and sublay mesh reinforcement versus primary suture only in midline laparotomies (PRIMA): 2-year follow-up of a multicentre, double-blind, randomised controlled trial. The Lancet. 2017;390(10094):567–567. doi:10.1016/S0140-6736(17)31332-6

3. Bosanquet DC, Ansell J, Abdelrahman T, et al. Systematic Review and Meta-Regression of Factors Affecting Midline Incisional Hernia Rates: Analysis of 14 618 Patients. Krieg A, ed. PLOS ONE. 2015;10(9):e138745. doi:10.1371/journal.pone.0138745

4. Fink C, Baumann P, Wente MN, et al. Incisional hernia rate 3 years after midline laparotomy. The British Journal of Surgery. 2014;101(2):51–51. doi:10.1002/bjs.9364

5. Van Ramshorst GH, Eker HH, Hop WC, Jeekel J, Lange JF. Impact of incisional hernia on health-related quality of life and body image: a prospective cohort study. The American Journal of Surgery. 2012;204(2):144–144. doi:10.1016/j.amjsurg.2012.01.012

6. Schlosser KA, Renshaw SM, Tamer RM, Strassels SA, Poulose BK. Ventral hernia repair: an increasing burden affecting abdominal core health. Hernia: The Journal of Hernias and Abdominal Wall Surgery. 2023;27(2):415–415. doi:10.1007/s10029-022-02707-6

7. Poulose BK, Shelton J, Phillips S, et al. Epidemiology and cost of ventral hernia repair: making the case for hernia research. Hernia. 2012;16(2):179–179. doi:10.1007/s10029-011-0879-9

8. Deerenberg EB, Harlaar JJ, Steyerberg EW, et al. Small bites versus large bites for closure of abdominal midline incisions (STITCH): a double-blind, multicentre, randomised controlled trial. Lancet. 2015;386(10000):1254–1254. doi:10.1016/S0140-6736(15)60459-7

9. van ‘t Riet M, Steyerberg EW, Nellensteyn J, Bonjer HJ, Jeekel J. Meta-analysis of techniques for closure of midline abdominal incisions. The British Journal of Surgery. 2002;89(11):1350–1350. doi:10.1046/j.1365-2168.2002.02258.x

10. Fortelny RH, Hofmann A, Baumann P, et al. Three-year follow-up analysis of the short-stitch versus long-stitch technique for elective midline abdominal closure randomized-controlled (ESTOIH) trial. Hernia: The Journal of Hernias and Abdominal Wall Surgery. 2024;28(4):1283–1283. doi:10.1007/s10029-024-03025-9

11. Fortelny RH, Andrade D, Schirren M, et al. Effects of the short stitch technique for midline abdominal closure on incisional hernia (ESTOIH): randomized clinical trial. The British Journal of Surgery. 2022;109(9):839–839. doi:10.1093/bjs/znac194

12. García-Ureña MÁ, López-Monclús J, Hernando LAB, et al. Randomized controlled trial of the use of a large-pore polypropylene mesh to prevent incisional hernia in colorectal surgery. Annals of Surgery. 2015;261(5):876–876. doi:10.1097/SLA.0000000000001116

13. Kokotovic D, Bisgaard T, Helgstrand F. Long-term Recurrence and Complications Associated With Elective Incisional Hernia Repair. JAMA. 2016;316(15):1575. doi:10.1001/jama.2016.15217

14. Ehlers AP, Sinamo JK, Howard R, et al. Long-Term Risk of Mesh Infection Requiring Removal After Elective Ventral Hernia Repair. JAMA Surgery. 2025;160(12):1318. doi:10.1001/jamasurg.2025.3915

15. Basta MN, Kozak GM, Broach RB, et al. Can We Predict Incisional Hernia?: Development of a Surgery-specific Decision–Support Interface. Annals of Surgery. 2019;270(3):544–544. doi:10.1097/SLA.0000000000003472

16. Fischer JP, Basta MN, Mirzabeigi MN, et al. A Risk Model and Cost Analysis of Incisional Hernia After Elective, Abdominal Surgery Based Upon 12,373 Cases: The Case for Targeted Prophylactic Intervention. Annals of Surgery. 2016;263(5):1010–1010. doi:10.1097/SLA.0000000000001394

17. Pereira-Rodríguez JA, Bravo-Salva A, Argudo-Aguirre N, Amador-Gil S, Pera-Román M. Defining High-Risk Patients Suitable for Incisional Hernia Prevention. Journal of abdominal wall surgery: JAWS. 2023;2:10899. doi:10.3389/jaws.2023.10899

18. Cherla DV, Moses ML, Mueck KM, et al. External Validation of the HERNIAscore: An Observational Study. Journal of the American College of Surgeons. 2017;225(3):428–428. doi:10.1016/j.jamcollsurg.2017.05.010

19. Goodenough CJ, Ko TC, Kao LS, et al. Development and validation of a risk stratification score for ventral incisional hernia after abdominal surgery: hernia expectation rates in intra-abdominal surgery (the HERNIA Project). Journal of the American College of Surgeons. 2015;220(4):405–405. doi:10.1016/j.jamcollsurg.2014.12.027

20. DePaolo J, Biagetti G, Judy R, et al. Polygenic Scoring for Detection of Ascending Thoracic Aortic Dilation. Circulation: Genomic and Precision Medicine. 2024;17(5). doi:10.1161/CIRCGEN.123.004512

21. DePaolo J, Zamirpour S, Abramowitz S, et al. Predicting Thoracic Aortic Dissection in a Diverse Biobank Using a Polygenic Risk Score. JACC: Advances. 2025;4(5):101743. doi:10.1016/j.jacadv.2025.101743

22. Privé F, Aschard H, Carmi S, et al. Portability of 245 polygenic scores when derived from the UK Biobank and applied to 9 ancestry groups from the same cohort. American Journal of Human Genetics. 2022;109(1):12–12. doi:10.1016/j.ajhg.2021.11.008

23. Inouye M, Abraham G, Nelson CP, et al. Genomic Risk Prediction of Coronary Artery Disease in 480,000 Adults. Journal of the American College of Cardiology. 2018;72(16):1883–1883. doi:10.1016/j.jacc.2018.07.079

24. Khera AV, Chaffin M, Aragam KG, et al. Genome-wide polygenic scores for common diseases identify individuals with risk equivalent to monogenic mutations. Nature Genetics. 2018;50(9):1219–1219. doi:10.1038/s41588-018-0183-z

25. Pregnall AM, Yuan S, Lawrence JM, et al. Decoding the genetic architecture of hernia through genome-wide association and multi-trait analyses. medRxiv. 2026;. doi:10.64898/2026.05.12.26353033

26. Ruan Y, Lin YF, Feng YCA, et al. Improving polygenic prediction in ancestrally diverse populations. Nature Genetics. 2022;54(5):573–573. doi:10.1038/s41588-022-01054-7

27. Verma A, Damrauer SM, Naseer N, et al. The Penn Medicine BioBank: Towards a Genomics-Enabled Learning Healthcare System to Accelerate Precision Medicine in a Diverse Population. Journal of Personalized Medicine. 2022;12(12):1974. doi:10.3390/jpm12121974

28. Lambert SA, Wingfield B, Gibson JT, et al. Enhancing the Polygenic Score Catalog with tools for score calculation and ancestry normalization. Nature Genetics. 2024;56(10):1989–1989. doi:10.1038/s41588-024-01937-x

29. Tanigawa Y, Qian J, Venkataraman G, et al. Significant sparse polygenic risk scores across 813 traits in UK Biobank. Ripatti S, ed. PLOS Genetics. 2022;18(3):e1010105. doi:10.1371/journal.pgen.1010105

30. Jung H, Jung HU, Baek EJ, et al. Integration of risk factor polygenic risk score with disease polygenic risk score for disease prediction. Communications Biology. 2024;7(1):180. doi:10.1038/s42003-024-05874-7

31. Pepe MS. Limitations of the Odds Ratio in Gauging the Performance of a Diagnostic, Prognostic, or Screening Marker. American Journal of Epidemiology. 2004;159(9):882–882. doi:10.1093/aje/kwh101

32. Wald NJ, Old R. The illusion of polygenic disease risk prediction. Genetics in Medicine. 2019;21(8):1705–1705. doi:10.1038/s41436-018-0418-5

33. Talwar AA, Desai AA, McAuliffe PB, et al. Optimal computed tomography-based biomarkers for prediction of incisional hernia formation. Hernia. 2023;28(1):17–17. doi:10.1007/s10029-023-02835-7

34. Elliott J, Bodinier B, Bond TA, et al. Predictive Accuracy of a Polygenic Risk Score–Enhanced Prediction Model vs a Clinical Risk Score for Coronary Artery Disease. JAMA. 2020;323(7):636. doi:10.1001/jama.2019.22241

35. Mosley JD, Gupta DK, Tan J, et al. Predictive Accuracy of a Polygenic Risk Score Compared With a Clinical Risk Score for Incident Coronary Heart Disease. JAMA. 2020;323(7):627. doi:10.1001/jama.2019.21782

36. Hornick MM, Talwar A, Voytik M, et al. Surgeon-Informed Clinical Decision Support Software for Surgical Risk Prediction and Outcomes Tracking. Journal of Surgical Research. 2025;315:937–949. doi:10.1016/j.jss.2025.09.089

37. Wei J, Attaar M, Shi Z, et al. Identification of fifty-seven novel loci for abdominal wall hernia development and their biological and clinical implications: results from the UK Biobank. Hernia: The Journal of Hernias and Abdominal Wall Surgery. 2022;26(1):335–335. doi:10.1007/s10029-021-02450-4

38. Muse ED, Chen SF, Liu S, et al. Impact of polygenic risk communication: an observational mobile application-based coronary artery disease study. npj Digital Medicine. 2022;5(1):30. doi:10.1038/s41746-022-00578-w

